## Supplementary figures and images for "Human and bacterial genetic variation shape oral microbiomes and health"

### Extended Data Table 1

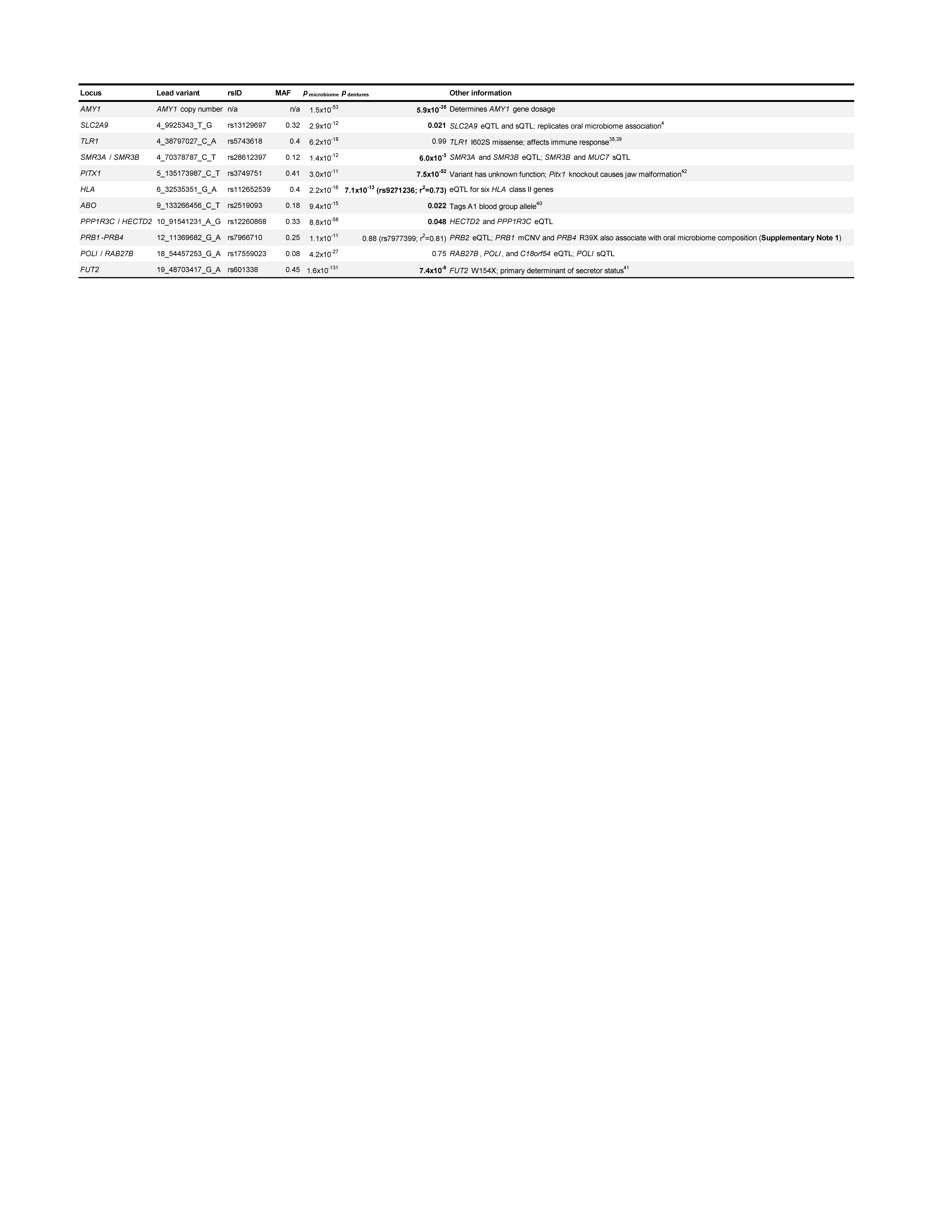
