## Supplementary Notes 1 through 8 and Supplementary Figures 1 through 3 for "Human and bacterial genetic variation shape oral microbiomes and health"

### Supplementary Note 1 – Complex variation at the salivary proline-rich protein locus

We characterized genetic associations with oral microbiome composition at the cluster of *PRB* genes on chromosome 12. As previous work had observed copy number variation here^1-3^, we wondered whether this could explain a substantial amount of the association signal. We estimated *PRB1* copy number in SPARK using whole genome sequencing reads that mapped to a 28.7kb region in GRCh38 previously seen to be copy number variable^3^ (chr12:11362789-11391498). Read depth was then normalized against the total number of reads that aligned in either of the 0.5Mb regions flanking this region.

SPARK participants were found to have a range of 0 to 7 diploid copies of *PRB1* (**Extended Data Fig. 2a**) and minimal copy number variation of the nearby homologous *PRB* genes (data not shown). Although *PRB1* copy number appeared to generate the strongest association for the species whose abundance best associated at this locus, *Stomatobaculum* SGB5266 (*p*=1.1x10^-13^, **Extended Data Fig. 2b**) and associated linearly (**Extended Data Fig. 2c**), the pattern of associations at the *PRB* locus with oral microbiome composition (from our mPC-based test) appeared to be more complex (*p*=6.7x10^-5^ for *PRB1* copy number, **Extended Data Fig. 2d**). A common loss-of-function variant in the nearby *PRB4* gene that associated strongly with the oral microbiome composition (rs12829245, *p*=3.3x10^-7^, **Extended Data Fig. 2d**) only nominally associated with *Stomatobaculum* SGB5266 abundance in a joint model with *PRB1* copy number (*p*=0.036, **Extended Data Fig. 2e**). Future work with larger sample sizes may allow for improved fine-mapping of genetic effects in the *PRB* locus and identification of species underlying the association pattern seen for oral microbiome composition.

1. Azen, E. A., O’Connell, P. & Kim, H. S. PRB2/1 fusion gene: a product of unequal and homologous crossing-over between proline-rich protein (PRP) genes PRB1 and PRB2. *Am. J. Hum. Genet.* **50**, 842–851 (1992).

2. Azen, E. A., Latreille, P. & Niece, R. L. PRBI gene variants coding for length and null polymorphisms among human salivary Ps, PmF, PmS, and Pe proline-rich proteins (PRPs). *Am. J. Hum. Genet.* **53**, 264–278 (1993).

3. Handsaker, R. E. *et al.* Large multiallelic copy number variations in humans. *Nat. Genet.* **47**, 296–303 (2015).

### Supplementary Note 2 – Replication of previous oral microbiome associations with *AMY1* copy number

In this note we examine the extent to which previously-reported associations of *AMY1* copy number with oral microbial abundance phenotypes (reported by Poole et al.^1^ and Hasegawa et al.^2^) replicated in the SPARK WGS cohort.

Poole et al.^1^ reported several associations between *AMY1* copy number and OTUs in the Greengenes 16S rRNA Gene Database (August 2013 release):

https://ftp.microbio.me/greengenes_release/gg_13_8_otus/taxonomy/99_otu_taxonomy.txt

The Greengenes database provides an approximate taxonomic mapping of each OTU (based on hierarchical clustering of sequences available at the time of release). These taxonomic classifications typically end at the genus level, such that it was not immediately clear how to map OTUs to species whose abundances we measured in SPARK using MetaPhlAn.

To obtain higher-resolution taxonomic classifications of OTUs from Greengenes (making use of microbial sequences that have been generated in the decade since the August 2013 release), we obtained their 16S rRNA sequences (available at <https://ftp.microbio.me/greengenes_release/gg_13_8_otus/rep_set/99_otus.fasta>) and submitted them to NCBI BLAST using the "rRNA/ITS databases" option. This approach produced the following results:

**OTU 4321396 “*Prevotella* (*Prevotellaceae*)”** clearly mapped to *Prevotella pallens*, with the top two matches at 97% identity for different *Prevotella pallens* accessions:

| **Description** | **Max Score** | **Query Cover** | **Per. ident** | **Acc. Len** | **Accession** |
| --- | --- | --- | --- | --- | --- |
| Prevotella pallens ATCC 700821 strain JCM 11140 16S ribosomal RNA, partial sequence | 2298 | 100% | 97.02 | 1492 | NR_113121.1 |
| Prevotella pallens strain 10371 16S ribosomal RNA, partial sequence | 2279 | 100% | 96.73 | 1454 | NR_026417.1 |
| Prevotella aurantiaca strain OMA31 16S ribosomal RNA, partial sequence | 2054 | 100% | 93.82 | 1481 | NR_112878.1 |
| Prevotella intermedia strain B422 16S ribosomal RNA, partial sequence | 1917 | 100% | 91.94 | 1459 | NR_026119.1 |
| Prevotella intermedia strain JCM 12248 16S ribosomal RNA, partial sequence | 1916 | 100% | 91.94 | 1494 | NR_113106.1 |

The effect direction of the association reported by Poole et al. agreed with the effect direction in SPARK, in which higher *AMY1* copy number associated with higher *Prevotella pallens* abundance.

**OTU 269907 “*Prevotella* (*Paraprevotellaceae*)”** mapped to various *Prevotella*-like or *Bacteriorides* species:

| **Description** | **Max Score** | **Query Cover** | **Per. ident** | **Acc. Len** | **Accession** |
| --- | --- | --- | --- | --- | --- |
| Prevotellamassilia timonensis strain Marseille-P2831 16S ribosomal RNA, partial sequence | 1792 | 100% | 90.52 | 1493 | NR_144750.1 |
| Alloprevotella rava strain 81/4-12 16S ribosomal RNA, partial sequence | 1766 | 99% | 90.25 | 1455 | NR_118334.1 |
| Pseudoprevotella muciniphila strain E39 16S ribosomal RNA, partial sequence | 1657 | 99% | 88.77 | 1476 | NR_173659.1 |
| Bacteroides faecichinchillae JCM 17102 16S ribosomal RNA, partial sequence | 1615 | 100% | 88.22 | 1486 | NR_113206.1 |
| Bacteroides clarus YIT 12056 16S ribosomal RNA, partial sequence | 1598 | 99% | 88.14 | 1479 | NR_112893.1 |

The low sequence identity with any of these sequences suggested that OTU 269907 might correspond to a species not represented in the 16S rRNA BLAST database. We therefore looked more closely at the 31 species whose abundances in SPARK associated with *AMY1* copy number at FDR<0.05 and noticed that *Alloprevotella* sp. Lung230 (*p*=0.00045; effect direction concordant with OTU 269907) was not present in the BLAST database and could potentially be a match. We retrieved its 16S rRNA sequence from the NCBI reference annotations (at contig position NZ_JACSUD010000006.1:1271-2798) and observed that it matched the OTU 269907 sequence more closely than any of the 16S rRNA sequences in the NCBI BLAST database (94% sequence identity). Thus it seems likely that OTU 269907 and *Alloprevotella* sp. Lung230 represent the same or similar entities.

**OTU 4465803 “*Porphyromonas*”** mapped fairly well to *Porphyromonas pasteri*:

| **Description** | **Max Score** | **Query Cover** | **Per. ident** | **Acc. Len** | **Accession** |
| --- | --- | --- | --- | --- | --- |
| Porphyromonas pasteri strain KUFDS01 16S ribosomal RNA, partial sequence | 2071 | 100% | 93.5 | 1491 | NR_136788.1 |
| Porphyromonas catoniae ATCC 51270 strain JCM 13863 16S ribosomal RNA, partial sequence | 2021 | 100% | 92.87 | 1451 | NR_113082.1 |
| Porphyromonas catoniae ATCC 51270 16S ribosomal RNA, partial sequence | 1932 | 98% | 91.82 | 1470 | NR_026230.1 |
| Porphyromonas loveana strain UQD444 16S ribosomal RNA, partial sequence | 1724 | 100% | 89.03 | 1492 | NR_152035.1 |
| Porphyromonas gulae strain JCM 13865 16S ribosomal RNA, partial sequence | 1714 | 99% | 89.11 | 1481 | NR_113088.1 |

In SPARK, we observed a nominally significant association with *AMY1* for this *Porphyromonas pasteri* (*p*=0.032) but the effect direction was opposite (lower abundance with high *AMY1* copy number).

**OTU 92430 “*Lachnospiraceae*”** did not map well to any species in the BLAST database:

| **Description** | **Max Score** | **Query Cover** | **Per. ident** | **Acc. Len** | **Accession** |
| --- | --- | --- | --- | --- | --- |
| Anaerotalea alkaliphila strain F-3ap 16S ribosomal RNA, partial sequence | 1821 | 99% | 88.43 | 1517 | NR_173597.1 |
| Abyssivirga alkaniphila strain L81 16S ribosomal RNA, partial sequence | 1744 | 100% | 87.43 | 1520 | NR_148837.1 |
| Petrocella atlantisensis strain 70B-A 16S ribosomal RNA, partial sequence | 1720 | 97% | 87.61 | 1490 | NR_164620.1 |
| Faecalicatena contorta strain DSM 3982 16S ribosomal RNA, partial sequence | 1716 | 99% | 87.12 | 1524 | NR_117147.1 |
| Faecalicatena contorta strain DSM 3982 16S ribosomal RNA, partial sequence | 1716 | 99% | 87.16 | 1521 | NR_104803.1 |

While we did observe associations of *AMY1* copy number with abundances of several species in the *Lachnospiraceae* family (such as *Stomatobaculum* SGB5266, *p*=1.2e-19), the effects directions were opposite for each of these (lower abundance with high *AMY1* copy number) except for *Lachnoanaerobaculum* sp. ICM7 (p=0.00027). However, the 16 rRNA sequence for *Lachnoanaerobaculum* sp. ICM7 has low homology with OTU 92430 (85.09% sequence identity), suggesting they are unlikely to represent the same entity.

Other *AMY1* copy number associations reported by Poole et al. clearly did not map to any associations we observed in SPARK: we did not identify associations with any species in the *Haemophilus* genus (for OTU 4406393), the *Neisseria* genus (for OTU 1106060), the *Leptotrichiaceae* family (for OTU 876114), the *Weeksellaceae* family (for OTU 968873), or to *Porphyromonas endodontalis* (for OTU 4423790).

Hasegawa et al.^2^ reported an association between *AMY1* copy number and abundance of the *Capnocytophaga* genus. We did not observe such a genus-level association in SPARK (p=0.49), but we did observe an association of *AMY1* copy number with abundance of one *Capnocytophaga* species (*Capnocytophaga* SGB2480 *p*=0.00021), such that it could be possible that the *Capnocytophaga* genus abundance measured by Hasegawa et al. actually maps onto a subset of *Capnocytophaga*.

In summary, we were able to replicate the two associations of *AMY1* copy number with *Prevotella* OTUs reported by Poole et al., are uncertain about the *Capnocytophaga* association seen in Hasegawa et al., and are confident that we did not replicate any of the other associations reported by Poole et al.

1. Poole, A. C. *et al.* Human Salivary Amylase Gene Copy Number Impacts Oral and Gut Microbiomes. *Cell Host Microbe* **25**, 553-564.e7 (2019).

2. Hasegawa, T. *et al.* Impact of salivary and pancreatic amylase gene copy numbers on diabetes, obesity, and functional profiles of microbiome in Northern Japanese population. *Sci. Rep.* **12**, 7628 (2022).

### Supplementary Note 3 – Functional assays of *AMY1* missense variants F141C and C477R

To investigate the molecular effects of these missense variants, we transfected HEK293T cells with plasmids carrying the reference coding sequence of *AMY1* or sequences encoding F141C and C477R mutant isoforms. Amylase protein expression was analyzed via western blot in both the supernatants and cellular lysates from each transfected line and subsequently purified for downstream enzymatic assays (see **Methods**). Cells with the reference sequence and the F141C sequence secreted amylase protein at similar levels, but cells with the C477R sequence did not; instead, the C477R amylase isoform appeared to be more glycosylated and retained intracellularly (**Extended Data Fig. 7a,b**). The F141C and reference amylase isoforms appeared to have equivalent enzymatic activity (t-test *p*=0.41, n=32 replicates of a fluorescent starch-degrading assay on purified secreted amylase; **Extended Data Fig. 7b,c**), suggesting that the F141C gain-of-function effect involves another mechanism, perhaps protein binding or activity on another substrate. Whether the retention and glycosylation of the C477R isoform *in vitro* is relevant to oral phenotypes remains to be determined.

### Supplementary Note 4 – Pathway relative abundance associations

At most of the 11 loci associated with oral microbial composition, human genetic variants associated with abundances of many microbial taxa (**Supplementary Table 2**). This led us to wonder whether the microbial species associated with a given locus might be united by shared utilization of biochemical pathways. To explore this question, we tested genetic variants for association with pathway abundance phenotypes that aggregate sequencing depth-of-coverage measurements across gene families within a pathway (and across species) using the HUMAnN pipeline^1^, following previous work^2^.

In more detail, an inclusive cohort-wide taxonomic profile was first generated by merging all metagenome relative abundance profiles (generated by MetaPhlAn) across the full SPARK WGS cohort using merge_metaphlan_tables.py before taking the max of each taxonomic entry across individuals with humann_reduce_table. An initial run of HUMAnN (v3.8) was then performed using this cohort taxonomic profile to produce a custom indexed reference of microbial coding sequences from taxa that were observed in any SPARK saliva sample. This custom reference was then used with subsequent HUMAnN runs for each sample, bypassing translated search of sequencing reads given that prior characterization of the healthy human oral microbiome should allow for high rates of gene and pathway assignments^1^. To avoid exceeding storage available on compute nodes as a consequence of sample input size, we implemented two filters to the unmapped reads used as input: 1) only the first read from each pair was taken, as HUMAnN does not use paired read information and mate reads are likely to be redundant; and 2) samples with >100 million reads were downsampled to ~100 million randomly selected reads. Output abundances were normalized to relative abundances before being merged across samples with humann_join_tables.

To test pathway abundances for association with human genetic variants, we performed rank-based inverse normal transformation (to avoid test statistic inflation from outlier values) on the relative abundance measures for each of the 613 observed pathways. These pathways were then tested for association with host genotypes using BOLT-LMM. The top 20 principal components from PCA on 338 pathways observed at >10% prevalence were used as covariates to control for the largest axes of variation across samples, along with sequencing batch, age, age squared, square root of age, sex, percent of mapped reads, and the top 10 genetic ancestry principal components.

Across the 11 loci identified from association analyses of oral microbiome composition, the abundance of 140 pathways (FDR<0.05) were significantly associated with genotypes we had found to associate with microbial abundances. These included an association between decreased utilization of the tricarboxylic acid (TCA) cycle and increasing *AMY1* copy number, which might indicate a shift to fermentation in individuals with more copies of the salivary amylase gene (*p*=6.2x10^-6^, **Extended Data Fig. 8a**, **Extended Data Table 2**).

Interestingly, upon inspecting pathways and gene families within species whose abundances associated with human genetic variants, we noticed that across gene family abundances for *Haemophilus sputorum*—the species whose abundance most strongly associated with *FUT2* loss-of-function—there was a single outlier which associated much more strongly with *FUT2* genotype than the coverage of other gene families (**Extended Data Fig. 8b**). This outlier appeared to correspond to a protein annotated as a trimeric autotransporter adhesin (Ata). We were curious whether this might be explained by variation within the *H. sputorum* genomes that associated with host *FUT2* genotype, motivating our subsequent search for bacterial gene dosages associated with human genetic variation, similar to a recent observation of such an effect in the gut microbiome^3^.

1. Beghini, F. *et al*. Integrating taxonomic, functional, and strain-level profiling of diverse microbial communities with bioBakery 3. *eLife* **10**, e65088 (2021).

2. Lopera-Maya, Esteban A., *et al*. Effect of host genetics on the gut microbiome in 7,738 participants of the Dutch Microbiome Project. *Nature Genetics* **54**, 143-151 (2022).

3. Zhernakova, D. V. *et al.* Host genetic regulation of human gut microbial structural variation. *Nature* **625**, 813–821 (2024).

### Supplementary Note 5 – Testing human genetic variants for association with microbial gene dosages

**Comparing and contrasting genetic association analysis of microbial abundance phenotypes versus microbial gene dosage phenotypes**

The key conceptual difference between testing human genetic variants for association with microbial abundances (**Fig. 2a**) versus microbial gene dosages (**Fig. 4a**) is that the latter approach searches for effects on relative fitness of microbial strains that do or do not contain a genomic region, whereas the former approach searches for effects on relative fitness of a microbial species. The main strength of the gene dosage-based approach is that it can simultaneously implicate both a human genetic variant and a microbial gene that are involved in a host-microbe genetic interaction, potentially providing insight into the molecular mechanism that underlies the interaction. However, the microbial abundance-based approach may be preferable for other purposes, such as considering the overall diversity and physical structure of the microbiome (or any property conferred by microbes rather than genes) or identifying non-commensal species that could potentially be therapeutic targets (based on colocalization of genetic associations with species abundances and oral health phenotypes).

The two approaches are somewhat complementary, as human genetic variants could associate with relative microbial abundances but not with microbial gene dosages or vice versa. For example, relative abundance phenotypes (which are based on counts of read alignments across many segments of a microbial genome) have less sampling noise than gene dosage phenotypes based on read alignments within a single genomic region (here 500bp), potentially offering better power to detect genetic associations. Additionally, a microbial species may not have many common structural variants that overlap genes that interact with human genotypes. On the other hand, a host genotype may strongly influence presence or absence of a microbial gene, yet have little or no detectable effect on abundance of the species due to factors such as (i) abundance having low variance (e.g., constrained by size of a niche) and/or (ii) abundance being primarily environmentally-driven. In either of these settings, dosage of a genomic region can still associate strongly with host genotype because the gene dosage phenotype controls for such environmental constraints or influences by comparing abundances of bacteria with and without the gene, in a manner analogous to how analyzing allele-specific expression can increase power to detect associations between genetic variants and gene expression (i.e., cis-eQTLs).

Beyond the above considerations, we also note that mismapping of sequencing reads is a greater concern for gene dosage analyses: whereas microbial abundance phenotypes are typically generated by counting alignments of sequencing reads to marker genes that uniquely specify taxa, the microbial gene dosage phenotypes that we generated and analyzed in this work considered all 500bp genomic regions of a reference panel of 30 bacterial genomes. As such, we took several precautions to mitigate potential effects of alignment artifacts, discussed in the remainder of this note.

**Technical subtleties of testing human genetic variants for association with bacterial gene dosages**

As described briefly in **Methods**, in order to robustly test human genetic variants for association with bacterial gene dosage phenotypes (derived from normalized read-depth measurements within 500bp bins of bacterial genomes), we employed a series of transformations and adjustments to reduce the risk of false positives.

First, we normalized a given sample’s read depth in each 500bp bin of a given bacterial genome by the median genomic coverage for that species in that sample (such that normalized read-depth measurements had a median of 1 among bins corresponding to each species). This controls for the differences in the abundance of the species across samples, avoiding associations with read-depth that reflect effects of a human genetic variant on species abundance rather than dosage of a specific gene. Normalizing using median coverage rather than mean coverage provides robustness to repetitive genomic regions with excessive coverage.

Second, we truncated these normalized read depths in each (bin)x(sample) to the range of [0,1]. The purpose of this was to focus on deletions that we expect to generate normalized coverages between 0 and 1 (depending on the fraction of bacteria with and without the deletion) and limit noise from samples that happen to have coverage >1 by chance. While this does not exclude the possibility of higher gene copy numbers contributing to an association, as higher copy numbers would still cause normalized read depth to saturate at 1 more often, the truncated read-depths are not expected to efficiently capture signals of copy-number-increasing variation; this is a limitation of our current approach.

The third adjustment we applied was the most substantial in what it attempted to control. This adjustment was to include, as covariates during association testing, principal components of the read-depth matrix (samples)x(truncated, median-normalized read depth bins for a given species). The reasoning here is that if there are multiple strains of a species circulating among human oral microbiomes, they will likely differ at multiple genomic regions that are present in some strains but not others, such that these regions will have normalized coverages that reflect the relative abundances of strains. If relative strain abundances are influenced by a human genetic variant, then normalized read-depth measurements in strain-informative regions will associate with the variant, but most of these associations will be non-specific in the sense that they do not point to dosages of genes that drive the association (and are thus uninformative for molecular interpretation). This phenomenon is similar to population stratification in conventional GWAS, in which ancestry-informative variants may be statistically correlated with a phenotype but biologically irrelevant. Consequently, just as genome-wide ancestry principal components are typically included as covariates in human genotype-phenotype association analyses, we included bacterial genomic coverage principal components as covariates when testing bacterial read-depth measurements for association with human genotypes. We decided to include 20 genomic coverage principal components to ensure control of any major strain effects (which should be captured by top principal components) while ensuring preservation of statistical power (as 20 is a small fraction of the SPARK sample size). However, we note that future approaches might optimize this parameter per species, given that the set of covarying genomic regions likely varies considerably across species.

Fourth, we controlled for any residual inflation of test statistics (from potential unknown confounders) by applying a form of genomic control^1^, again borrowing from techniques previously applied in genome-wide association studies for human variants. Specifically, we take the t-values from linear regression of each read depth bin across the genome of a species and square them to get test statistics that are approximately X_1_^2^-distributed. These are then divided by the quantity, median(X_1_^2^)/F^-1^(0.5), where median(X_1_^2^) is the median of X_1_^2^ statistics across bins in that genome and F^-1^(0.5) is the inverse cumulative distribution function for a X_1_^2^ random variable. In practice, inflation was fairly minimal, with a mean of 1.12 and standard deviation of 0.16 across 330 (species)x(human genotype) pairs.

Lastly, one concern that we recognize the above adjustments do not directly guard against is the possible scenario in which (i) mismapping occurs unevenly from another species B onto regions of species A’s genome, (ii) species A but not species B is in our reference panel, and (iii) the abundance of species B associates with a human genotype G. In this scenario, an association between genotype G and the abundance of species B could generate an apparent association of genotype G with gene dosage at one or more genomic bins of species A. We note that a large extent of mismapping from B onto A, as might occur with closely related species or strains, would yield multiple linked gene dosage associations that would likely be corrected for by the genomic coverage PCs in the third adjustment mentioned above. However, a focal mismapping onto a specific region (e.g., a region that was recently horizontally transferred to the bacterial isolate used to generate a given reference genome) would likely not be corrected.

While we cannot formally exclude the possibility of this last form of potential confounding, a few lines of evidence suggest that it is not a major contributor to the gene dosage associations that we identified. One reason is that if an association of genotype G with gene dosage in species A were due to mismapping from species B, then the abundance of species B should have a much more significant association with genotype G than the less-powered association of G with gene dosage in species A. That is, read alignments to a 500bp bin of species A (even if solely due to mismapping from species B) should provide a noisier quantification of species B’s abundance than the quantification computed by MetaPhlAn, which utilizes numerous marker genes throughout the genome of species B. On the contrary, the bacterial gene dosage associations we highlighted in this work had association test statistics exceeding all but the most significant associations of human genotypes with relative abundance phenotypes. Additionally, one would expect such mismapping-driven gene dosage associations to point to bacterial genes with functions unrelated to the human genetic variant, whereas for most key examples we highlighted, the associated regions contained genes that encode proteins with immediately plausible molecular relevance (glycoside hydrolase in *Prevotella*, amylase-binding proteins in *Streptococcus parasanguinis*, adhesins). In future analyses, expanding the panel of microbial reference genomes should decrease the potential of misalignment, though care will also be needed to handle potential multi-mapping of sequencing reads to regions of high homology across species.

**Rationale for testing 11 microbiome composition-associated human genetic variants for association with gene dosage (read-depth) phenotypes for 30 bacterial genomes**

Our approach of testing only 11 human genetic variants (nominated by our GWAS of microbiome composition) for association with gene dosages of only 30 bacteria has important limitations: for example, other human genetic loci may have stronger effects on microbial gene dosage but remain undetected, or other microbes (not included among the 30) may affect gene dosage through microbe-microbe interactions. We took this approach for two main reasons: (1) to minimize multiple hypothesis testing burden (by testing 11 variants rather than millions of common variants in the human genome); and (2) because identifying high-quality reference genomes for bacterial species required manual curation and often was not even possible. For example, many of the species genomic bins (SGB) quantified by the MetaPhlAn pipeline are derived from metagenomic bins and lack reference genomes. Pilot analyses running GWAS to test all common human genetic variants for association with exemplar gene dosage phenotypes did not detect strong associations at any loci beyond the 11 included in our tests (**Extended Data Fig. 9c-h**), suggesting that restricting to the 11 microbiome composition-associated lead variants did not greatly reduce discovery potential, but we anticipate that future analyses using new approaches that overcome these limitations will discover additional effects.

1. Devlin, B. & Roeder, K. Genomic control for association studies. *Biometrics* **55**, 997–1004 (1999).

### Supplementary Note 6 - *Streptococcal* amylase-binding protein is selected for in humans with higher *AMY1* copy number

Two genomic regions of *Streptococcus parasanguinis*, a common and generally benign member of dental plaque, associated with host *AMY1* copy number, and these two genomic regions overlapped its *abpA* and *abpB* (amylase-binding protein) orthologs^1,2^ (*p*=1.13x10^-7^ and 1.80x10^-7^, respectively; **Extended Data** **Fig. 8c**). The frequency at which these regions were present in oral microbiomes (based on normalized read-depth) increased with increasing host *AMY1* copy number, forming a non-linear allelic series with larger effects at lower copy numbers (**Extended Data Fig. 8d**). This relationship mirrors the higher abundance of both *abp* and *AMY1* gene copies in hominins relative to other primates^3^. These associations were specific to *abpA* and *abpB*; abundance of *S. parasanguinis* did not associate with *AMY1* copy number (**Extended Data Fig. 8e**). Both *S. parasanguinis* amylase binding proteins were computationally predicted by AlphaFold3^4^ to interact with the amylase protein face containing the F141C mutation (**Extended Data** **Fig. 8f,g** and **Supplementary Table 9**), suggesting that the F141C substitution may modulate interactions with amylase binding proteins to yield its effects on oral health phenotypes.

1. Gwynn, J. P. & Douglas, C. W. I. Comparison of amylase-binding proteins in oral streptococci. *FEMS Microbiol. Lett.* **124**, 373–379 (1994).

2. Scannapieco, F. A. Saliva-Bacterium Interactions in Oral Microbial Ecology. *Crit. Rev. Oral Biol. Med.* **5**, 203–248 (1994).

3. Fellows Yates, J. A. *et al.* The evolution and changing ecology of the African hominid oral microbiome. *Proc. Natl. Acad. Sci. U. S. A.* **118**, e2021655118 (2021).

4. Abramson, J. *et al.* Accurate structure prediction of biomolecular interactions with AlphaFold 3. *Nature* **630**, 493–500 (2024).

### Supplementary Note 7 – chr21 rRNA associations driven by dietary DNA co-acquisition

Prior to filtering the human genetic variants considered in our GWAS of oral microbiome composition to “well-behaved” regions of the human genome (based on inclusion in the TOPMed-r3 imputation reference panel), we observed two nearby GWAS peaks on the short arm of chromosome 21, with lead variants 21_8397578_T_C (2.50x10^-70^) and 21_8215817_C_G (4.90x10^-8^). Closer inspection revealed that these variant calls lay within copies of 28S rRNA genes, *RNA28SN3* and *RNA28SN2*, in the ribosomal RNA gene array of chromosome 21 at GRCh38 positions chr21:8396926-8401980 and chr21:8213888-8218941, respectively. Testing 21_8397578_T_C genotype calls for association with the abundances of individual microbial species showed that 21_8397578_T_C associated most strongly with *Streptococcus thermophilus* (p=8.2x10^-17^) and *Lactococcus cremoris* (p=1.8x10^-6^), species typically associated with fermented dairy production for foods such as cheese and yogurt. We therefore wondered whether the genotype calls for 21_8397578_T_C in SPARK might arise from a variant-calling artifact, reflecting presence of sequenced DNA originating from mammalian species involved in the dairy production process, such as cows, rather than human genetic variation.

Using BLAT^1^ to find the closest matches in the cow reference genome (ARS-UCD1.2/bosTau9) to the human *RNA28SN3* and *RNA28SN2* sequences identified the bosTau9 region chr27:6,217,392-6,221,526 as the best match for both human rDNA sequences, with complete coverage and 96.9% and 97% identity, respectively. Notably, the positions on the p-arm of chromosome 21 at which variant calls in the SPARK WGS data set most strongly associated with oral microbiome composition map to human-bovine rDNA sequence differences within stretches of high identity that likely allow for alignment of bovine DNA sequences to GRCh38 and subsequent (artifactual) variant calling:

| **Human**  **CHR** | **Human**  **POS** | **Human base** | **Bovine**  **CHR** | **Bovine POS** | **Bovine**  **base** | **DeepVariant call (SPARK WGS)** | **-log10(p)** |
| --- | --- | --- | --- | --- | --- | --- | --- |
| 21 | 8397578 | T | 27 | 6218050 | C | 21_8397578_T_C | 69.60 |
| 21 | 8397585 | A | 27 | 6218057 | G | 21_8397585_A_G | 67.22 |
| 21 | 8397564 | T | 27 | 6218036 | C | 21_8397564_T_C | 51.92 |
| 21 | 8397597 | C | 27 | 6218069 | G | 21_8397597_C_G | 43.95 |
| 21 | 8397419 | T | 27 | 6217885 | C | 21_8397419_T_C | 37.06 |
| 21 | 8397445 | C | 27 | 6217911 | T | 21_8397445_C_T | 26.42 |
| 21 | 8397409 | T | 27 | 6217875 | C | 21_8397409_T_C | 11.39 |
| 21 | 8397665 | C | 27 | 6218137 | C | 21_8397665_C_T | 10.16 |
| 21 | 8397407 | G | 27 | 6217873 | A | 21_8397407_G_A | 10.03 |
| 21 | 8215817 | C | 27 | 6219174 | C | 21_8215817_C_G | 7.31 |
| 21 | 8215840 | A | 27 | 6219197 | A | 21_8215840_A_C | 7.30 |

These lines of evidence suggest that the associations of variant calls in the rDNA locus of chromosome 21 with oral microbiome composition are driven by dietary co-acquisition of DNA from bacteria and cows (in dairy products) rather than true heritable variation. We hypothesize that some of the weaker associations, such as 21_8397665_C_T, that do not correspond to sequence differences with this region of the cow genome may be from other dairy animals (e.g., goat, sheep, water buffalo, yak) or to other rDNA repeats in the cow genome.

1. Kent, W. J. BLAT—The BLAST-Like Alignment Tool. *Genome Res.* **12**, 656–664 (2002).

### Supplementary Note 8 – Limitations of microbial gene dosage analyses

Our analytical approach for identifying host-microbe genetic interactions had several limitations. In particular, while limiting these analyses to 11 human genetic variants (those that already associated with oral microbiome composition) reduced multiple testing burden, this choice to improve statistical power to observe effects of these 11 loci came with the trade-off of potentially overlooking effects of human genetic variants at untested loci. Exploratory GWAS searching for effects of other variants on two sets of gene dosage phenotypes did not identify any additional strong associations at other loci in the human genome (**Extended Data Fig. 9c-h**). However, future analyses using an expanded set of microbial reference genomes and testing a broader set of human genetic variants are likely to identify additional effects, even within the SPARK data set**.** Another limitation of the gene dosage approach used here to identify potential genomic deletions based on reduced read-depth—in lieu of precisely calling microbial structural and sequence variants—is that the specific variation(s) in microbial genomes that drive observed statistical associations are unclear and require further characterization and validation. A reference panel of common structural variants in oral microbial species (generated from long-read sequencing data) could aid future analyses. Lastly, while we replicated many of the identified associations in another cohort (*All of Us*), analyses of cohorts representing other geographic regions could reveal novel interactions with environmental and dietary differences.

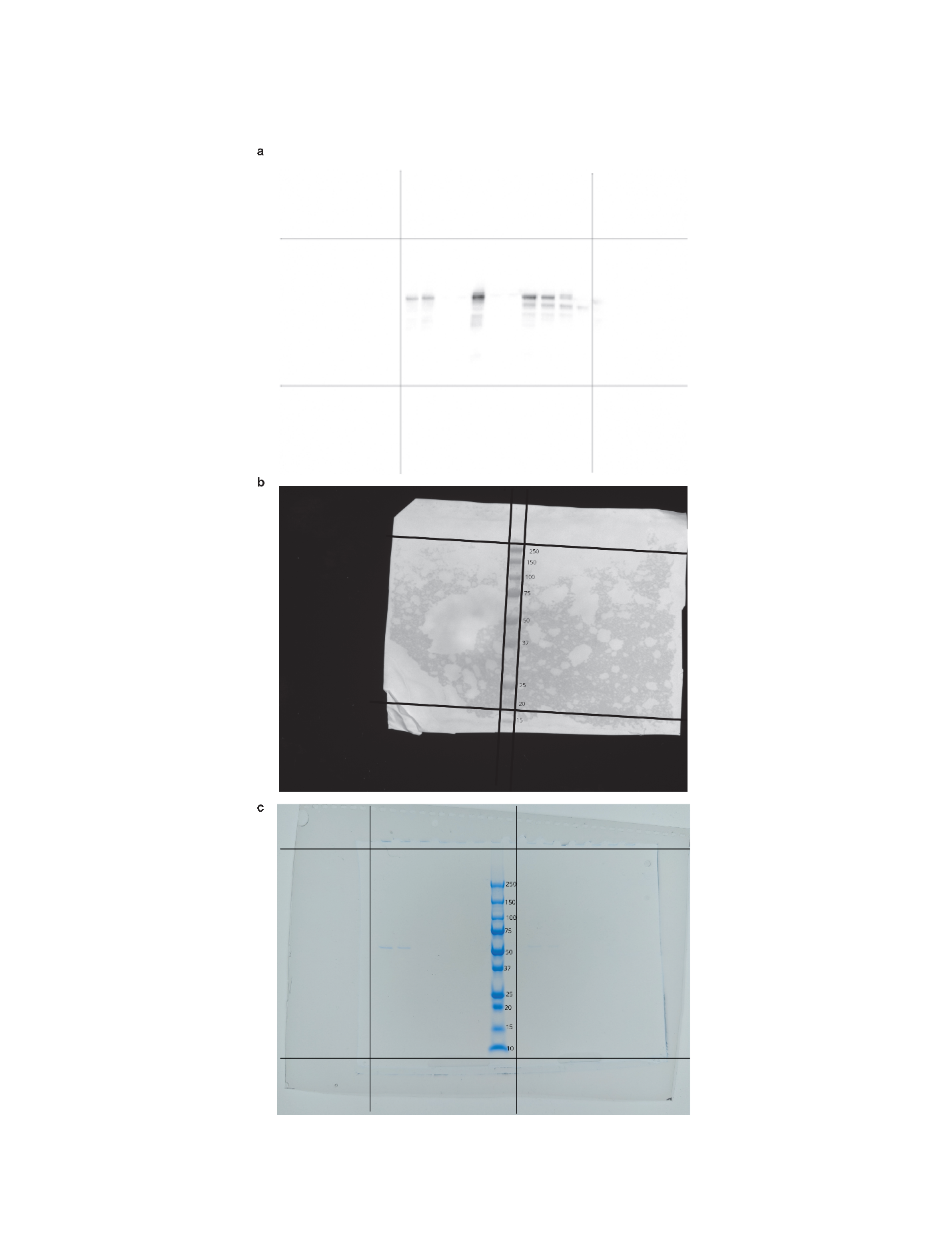

### Supplementary Figure 1. Source images for Extended Data Figure 7.

**a**, Raw chemiluminescence from Western blot for amylase transgenically expressed in HEK293T cells. **b**, Raw colorimetric image from the same Western blot in the same position as in a to image molecular weight ladder. **a** and **b** were cropped as indicated and superimposed to generate **Extended Data Figure 7a**. **c**, Source photograph taken of protein gel stained with Coomassie blue.

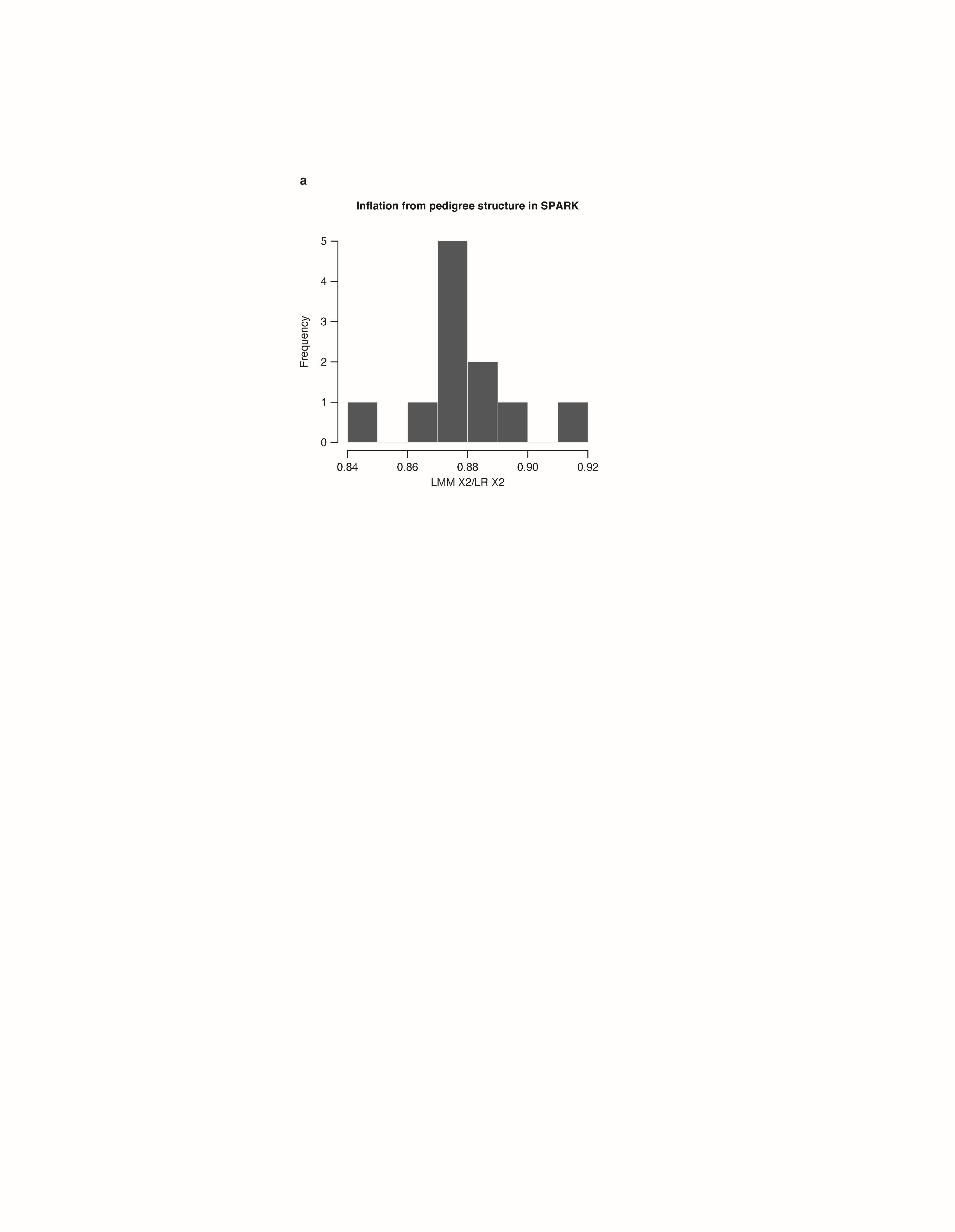

### Supplementary Figure 2. Modest inflation of chi-squared test statistics from linear regression in SPARK.

Chi-squared statistics from linear mixed models (LMM, which account for sample relatedness) are modestly lower than chi-squared statistics from linear regression (LR) for the 11 lead variants that associated with oral microbiome composition.

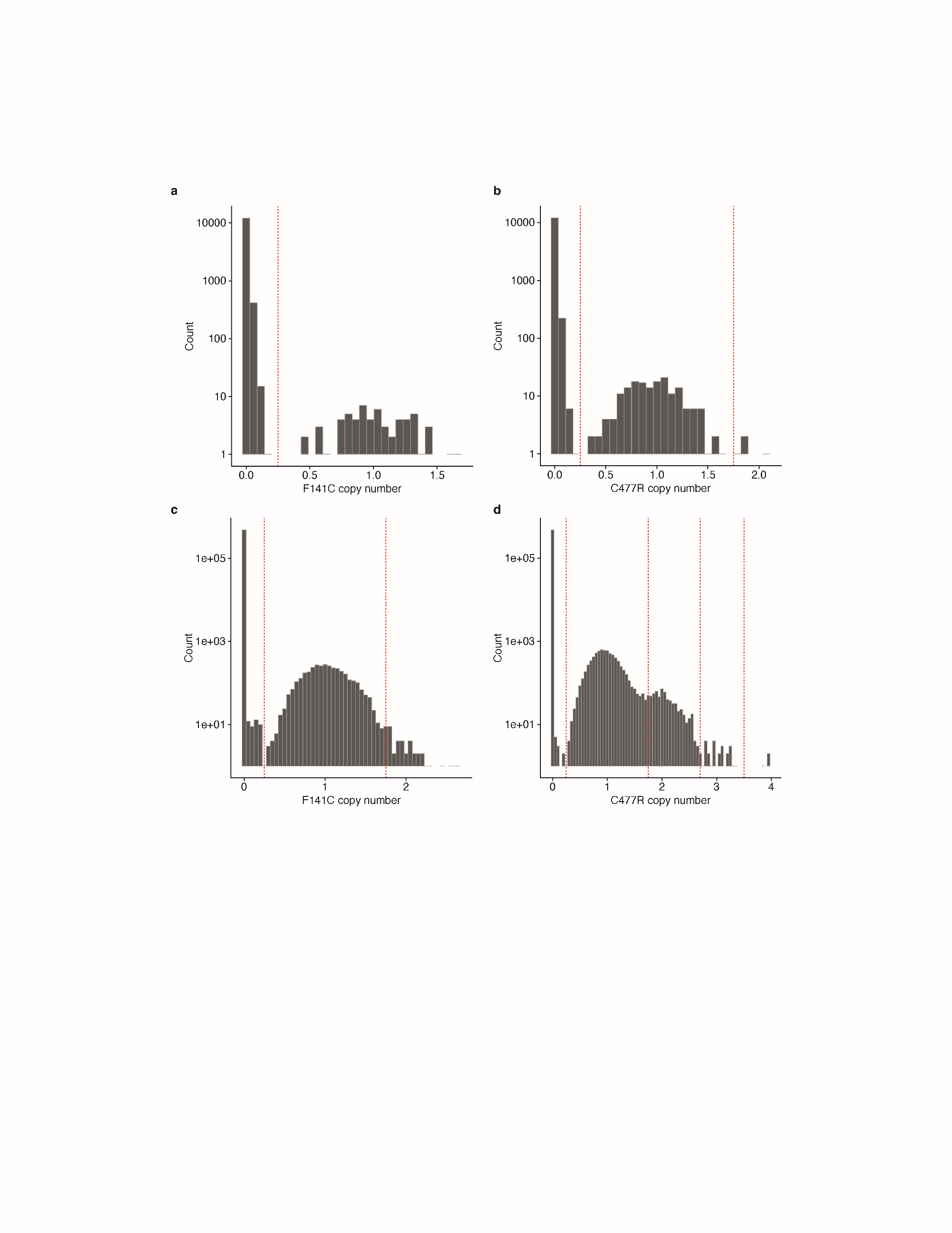

### Supplementary Figure 3. Copy number estimates of *AMY1* paralogous sequence variants in UK Biobank and SPARK.

**a**, Histograms of copy number estimates for *AMY1* F141C in SPARK (n=12,519). Thresholds used to call integer copy numbers are indicated with dotted red lines. **b**, Analogous to **a**, but for *AMY1* C477R. **c**, Analogous to **a**, but for UK Biobank (n=490,415). **d**, Analogous to **b**, but for UK Biobank.
